# Role of Cardiorespiratory Fitness and Mitochondrial Energetics in Reduced Walk Speed of Older Adults with Diabetes in the Study of Muscle, Mobility and Aging (SOMMA)

**DOI:** 10.1101/2023.11.03.23297992

**Authors:** Sofhia V. Ramos, Giovanna Distefano, Li-Yung Lui, Peggy M. Cawthon, Philip Kramer, Ian J. Sipula, Fiona M. Bello, Theresa Mau, Michael J. Jurczak, Anthony J. Molina, Erin E. Kershaw, David J. Marcinek, Frederico G.S. Toledo, Anne B. Newman, Russell T. Hepple, Stephen B. Kritchevsky, Bret H. Goodpaster, Steven R. Cummings, Paul M. Coen

## Abstract

**Rationale:** Cardiorespiratory fitness and mitochondrial energetics are associated with reduced walking speed in older adults. The impact of cardiorespiratory fitness and mitochondrial energetics on walking speed in older adults with diabetes has not been clearly defined.

**Objective:** To examine differences in cardiorespiratory fitness and skeletal muscle mitochondrial energetics between older adults with and without diabetes. We also assessed the contribution of cardiorespiratory fitness and skeletal muscle mitochondrial energetics to slower walking speed in older adults with diabetes.

**Findings:** Participants with diabetes had lower cardiorespiratory fitness and mitochondrial energetics when compared to those without diabetes, following adjustments for covariates including BMI, chronic comorbid health conditions, and physical activity. 4-m and 400-m walking speeds were slower in those with diabetes. Mitochondrial oxidative capacity alone or combined with cardiorespiratory fitness mediated ∼20-70% of the difference in walk speed between older adults with and without diabetes. Further adjustments of BMI and co-morbidities further explained the group differences in walk speed.

**Conclusions:** Skeletal muscle mitochondrial energetics and cardiorespiratory fitness contribute to slower walking speeds in older adults with diabetes. Cardiorespiratory fitness and mitochondrial energetics may be therapeutic targets to maintain or improve mobility in older adults with diabetes.

**ARTICLE HIGHLIGHTS:** Why did we undertake this study?

- To determine if mitochondrial energetics and cardiorespiratory fitness contribute to slower walking speed in older adults with diabetes.

What is the specific question(s) we wanted to answer?

- Are mitochondrial energetics and cardiorespiratory fitness in older adults with diabetes lower than those without diabetes? How does mitochondrial energetics and cardiorespiratory fitness impact walking speed in older adults with diabetes?

What did we find?

- Mitochondrial energetics and cardiorespiratory fitness were lower in older adults with diabetes compared to those without diabetes, and energetics, and cardiorespiratory fitness, contributed to slower walking speed in those with diabetes.

What are the implications of our findings?

- Cardiorespiratory fitness and mitochondrial energetics may be key therapeutic targets to maintain or improve mobility in older adults with diabetes.

**Graphical Abstract:** 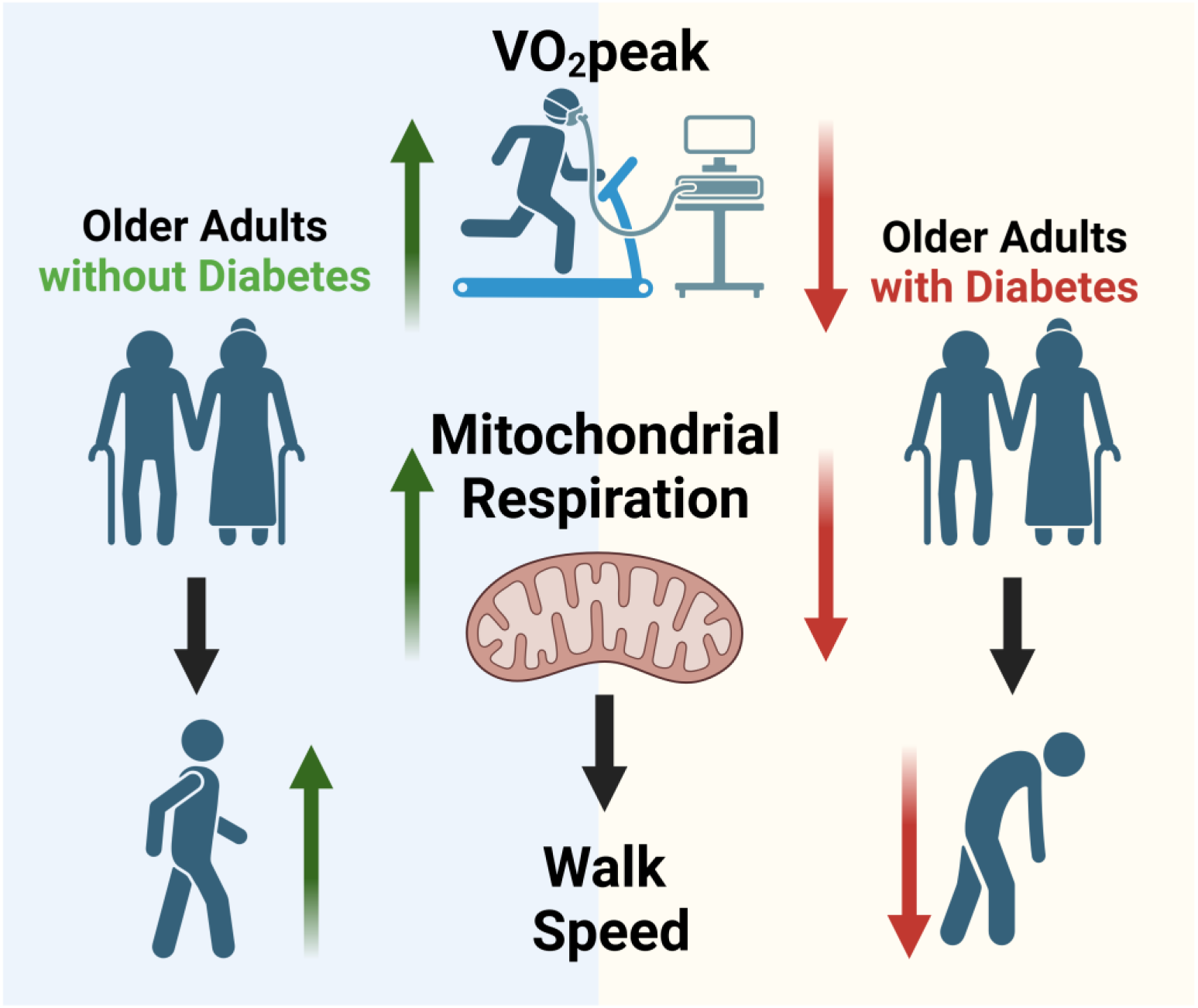

## Introduction

The US population is rapidly aging, and in 2022 the percentage of Americans aged 65 and older diagnosed with diabetes remains high at 30% [1]. This represents a healthcare challenge as older adults with diabetes have a greater risk of cardiovascular disease and mobility disability [2, 3]. The contributors to impaired mobility in older individuals with diabetes include the presence of co-morbidity, increased body mass index (BMI) [4], and lower muscle strength and quality [5–7]. However, the relationship between diabetes and loss of mobility remains only partially explained, and other contributors also likely play a role. Our group and others reported that cardiorespiratory fitness and skeletal muscle mitochondrial energetics are linked to reduced walking speed in aging, which is a predictor of mobility disability [8–10]. For example, muscle bioenergetics assessed using ^31^P-MRS was highly correlated with walking speed and partially explained age-related poorer performance in short (6-m) and long (400-m) walking tasks [10]. Several studies have found a close relationship between cardiorespiratory fitness and walking speed in older adults [11], which suggests that the decline in aerobic capacity also contributes to slower walking speed and mobility disability with age.

Muscle biopsies from individuals with diabetes have been shown to have lower oxidative phosphorylation (OXPHOS) capacity compared with muscle from healthy controls [12–14]. However, some studies [15, 16], but not all [12, 13, 17], indicate no differences between patients with diabetes and BMI-matched controls, indicating that obesity *per se* may underlie the lower muscle oxidative capacity. Indeed, other studies in people without diabetes have shown that BMI is strongly related to muscle oxidative capacity [18]. In addition, reports assessing mitochondrial energetics in older adults with diabetes tend to have small sample size and lack adjustment for critical confounding variables including physical activity and adiposity [13, 14, 17], and there are few reports focusing on older adults with diabetes specifically. Lower fitness is a predictor of insulin resistance and diabetes [19], and prevalence of diabetes is greater in individuals with low fitness [20]. It is unknown whether diabetes associated changes in cardiorespiratory fitness and/or muscle energetics explain slower walking speed seen in people with diabetes, independent of other covariates.

In this analysis, we leveraged data from the Study of Muscle, Mobility and Aging (SOMMA) [21], to investigate the association of diabetes with cardiorespiratory fitness and mitochondrial energetics in a cohort of 879 older adults who were well phenotyped in terms of the free living physical activity behaviors and body composition. We utilized ^31P^MRS, to measure muscle mitochondrial energetics *in vivo* (ATPmax) and high-resolution respirometry to measure mitochondrial energetics in muscle biopsies, a complementary approach that interrogates mitochondrial OXPHOS capacity at the myocellular level. Here, we tested the hypothesis that diabetes would be associated with lower cardiorespiratory fitness and muscle energetics independent of adiposity, physical activity levels, chronic comorbid health conditions, and oral hypoglycemic medication use. We also tested whether cardiorespiratory fitness and mitochondrial energetics mediate slower walking speed of older adults with diabetes.

## RESEARCH DESIGN AND METHODS

### Participant Recruitment

The Study of Muscle Mobility and Aging (SOMMA; http://sommaonline.ucsf.edu) is a prospective longitudinal cohort study of older adults and was designed to understand the biological basis of muscle aging [21]. Men and women (70+ yrs. old) were recruited between April 2019 to December 2021. Participants were enrolled at University of Pittsburgh and Wake Forest University School of Medicine if they were willing and able to complete a skeletal muscle biopsy and undergo Magnetic Resonance Imaging (MRI) and Spectroscopy (MRS). Exclusion criteria included an inability to walk one-quarter of a mile or climb a flight of stairs; had a BMI>40kg/m2; had an active malignancy or dementia; or any medical contraindication to biopsy or MR. Finally, participants must have been able to complete the 400-m walk test; those who appeared as they might not be able to complete the 400-m walk at the in-person screening visit completed a short distance walk (4-m) to ensure their walking speed was ≥ 0.6m/s. Presence of diabetes was identified by self-report, use of prescribed hypoglycemic medication, or having an HbA1c ≥6.5%. WIRB-Copernicus Group (WCG) Institutional Review Board (WCGIRB, study number 20180764) approved the study as the single IRB and all participants provided written informed consent.

### Baseline Assessments

Detail on study design and methodology have been published elsewhere [21]. The SOMMA Baseline Visit generally consisted of three days of assessments over several weeks: on “Day 1” the 400-m walk plus most other in-person assessments; “Day 2” cardiopulmonary exercise testing (CPET) and Magnetic Resonance Imaging (MRI) and Spectroscopy (MRS) were assessed; “Day 3” muscle biopsy specimen was collected. Age, sex, and race were collected by self-report. The self-reported medical conditions (heart disease, stroke, kidney disease/renal failure, and peripheral vascular disease) were assessed to determine if participants had those conditions or not. Participants were asked to bring all prescription medications they had taken in the 30 days prior to their “Day 1” clinic visit. If a participant forgot to bring one or more medications, clinic staff obtained this information over the telephone or at the return visit. The prescription medication use was reviewed and updated at the additional baseline visit days where CPET and tissue sampling were done. Body weight, height, and physical function assessments were also measured and accelerometry devices were provided. Most SOMMA participants completed cardiopulmonary exercise testing (CPET), magnetic resonance spectroscopy (MRS) and provided a percutaneous biopsy of the vastus lateralis muscle which was performed under fasted conditions. On the day of the muscle biopsy, participants prescribed oral hypoglycemic medication were asked to take their medications following tissue sampling.

### Mobility Assessments

Mobility was assessed by 400m and 4m walking tests. For the 400-m walk test, participants were instructed to walk 10 complete laps around a set course at their usual pace and without overexerting themselves. 4m walking speed was measured in 2 trials and the participant’s best time was used to calculate walking speed in m/sec.

### Physical Activity Assessments

Two devices were used to assess activity: the thigh-worn activPal that recorded total daily step count and the wrist-worn ActiGraph GT9x. Devices were placed on the participants at the baseline “Day 1” visit with a data collection period of 7 full days. Data was included only if the device was worn over a 24-hour period and had ≥17hour per day wear time. Activity levels collected from ActiGraph were determined by cut points described by Montoye and colleagues [22], and included total daily physical activity time, and time spent in sedentary, light, and moderate to vigorous physical activity (MVPA).

### Cardiopulmonary Exercise Testing Assessment

Participants walked for 5 minutes at a preferred walking speed, and progressive symptom-limited exercise protocol ensued with increases in speed (0.5 mph) and incline (2.5%) in 2-minute increments using a modified Balke protocol or a manual protocol [23, 24]. Cardiorespiratory fitness was determined as VO_2_peak (mL/min) which was identified as the highest 30-second average of VO_2_ (mL/min) achieved.

### Magnetic Resonance (MR) Imaging and Spectroscopy

^31^Phosphorous-MRS was used to assess *in-vivo* mitochondrial adenosine triphosphate (ATP) generation in the quadriceps muscle during an acute bout of knee isometric extension exercise [21, 24]. A 3 Tesla magnetic resonance magnet (Siemens Medical Systems – Prisma [Pittsburgh] or Skyra [Wake Forest]) with a 2.5” surface RF coil was used for the assessment. All data were analyzed using jMURI v7.0 [25]. Magnetic resonance imaging (MRI) was performed with a 2.5” surface RF coil and used to assess abdominal subcutaneous adipose tissue (ASAT), visceral adipose tissue (VAT), and quadricep muscle fat infiltration (QFI). The entire body was scanned and images were analyzed using AMRA Researcher® (AMARA medical AB, Linköping Sweden) [26].

### Muscle Biopsy and Mitochondrial Respiration

Muscle biopsy was completed with a Bergström trocar (5 or 6mm) as previously described [24]. Approximately 10 mg of muscle tissue was allocated for respirometry assays and were immediately submerged in ice cold BIOPS buffer. Fiber bundles of approximately 2-3 mg wet weight were gently teased apart and permeabilized as previously described [24] and weight was measured using an analytical balance (Mettler Toledo, Columbus, OH). Permeabilized fiber bundle (PmFB) oxygen consumption was measured using a high-resolution respirometer (Oxygraph-2k, Oroboros Instruments, Innsbruck, Austria). PmFB were placed into chambers containing MiR05 buffer supplemented with 25µM blebbistatin in the absence of light. Two protocols were completed in duplicate measuring carbohydrate supported (Protocol 1: 5mM pyruvate and 2mM malate) and fatty acid oxidation (FAO) supported (Protocol 2: 25µM palmitoyl-carnitine and 2mM malate) respiration. For protocol 1, state 3 respiration was stimulated with the addition of 4.2mM ADP (OXPHOS_CHO_) followed by subsequent additions of 10mM glutamate and 10mM succinate sequentially (maxOXPHOS_CHO_). Maximal uncoupled respiration was stimulated with consecutive 0.5uM titrations of 2-[2-[4-(trifluoromethoxy)phenyl] hydrazinylidene-propanedinitrile (FCCP) until maximum electron transport system (ETS) capacity was attained (maxETS_CHO_). For protocol 2, state 3 respiration was stimulated with the addition of 4mM ADP (OXPHOS_FAO_) followed by subsequent additions of 10mM glutamate and 10mM succinate to further stimulate mitochondrial complexes I and II (maxOXPHOS_FAO_). PmFB eliciting an oxygen consumption response <15% were not included in the final analysis. Respiratory control ratio (RCR) for protocol 1 (7.11 ± 2.86) and protocol 2 (3.47 ± 1.74) were assessed for quality control. Respiration was analyzed using DatLab 7.4.0.4. In SOMMA, not all participants completed both protocols due to time constraints. Protocol 1 (carbohydrates) was prioritized over Protocol 2 (fatty acids) when limited technician time was available, which left fewer participants with complete data for Protocol 2.

### Statistical Analysis

Differences in participant characteristics between older adults with and without diabetes were compared using the T-test for continuous variables (or Kruskal-Wallis non-parametric test for skewed variables) and chi-square test for dichotomized variables. Linear regressions without adjustment were performed to determine differences in cardiorespiratory fitness, and mitochondrial energetics between those with and without diabetes, and additional adjustments for the following confounders: age, race, gender, site/technician, BMI, and chronic conditions (model 1). Model 1 was further adjusted for steps per day from ActiPal or volume of adipose tissue depots: Abdominal Subcutaneous Adipose Tissue (ASAT), Visceral Adipose Tissue (VAT) and muscle Quadricep Fat Infiltration (QFI). Lastly, to determine the impact of hypoglycemic medication use within the group of older adults with diabetes, multivariate linear regression was performed adjusting for age, race, and site/technician. Those analyses were completed using JMP® Software, Version 16.

Linear regression was also used to determine the association of diabetes with walking speed. We evaluated the potential mediating influence of VO_2_peak and/or mitochondrial energetics. We did this by comparing the beta coefficient for the association of diabetes status on walking speed from the 400m and 4m walk. The base model included the confounding variables: gender, age, and race. model 2: base model plus technician and maxOXPHOS_CHO_, model 3: base model plus technician and maxOXPHOS_FAO_, model 4: base model plus clinical site and VO_2_peak, model 5: base model plus technician, VO_2_peak and maxOXPHOS_CHO_, model 6: base model plus technician, VO_2_peak and maxOXPHOS_FAO_. Fully adjusted models also included the potential mediators/confounders BMI, and chronic conditions. Finally, we compared the percent difference in the beta coefficient between the base model and subsequent models to understand how each adjustment impacted group differences in walking speed. Those analyses were performed using SAS version 9.4 (SAS institute Inc., Cary, NC).

## RESULTS

### Participant Characteristics

A total of 879 participants (59.2% women) with an average age of 76.5 ± 5.0 years were enrolled in SOMMA [21]. A summary of participants with available data are included in **Supplemental Figure 1.** Participant characteristics are summarized in **Table 1**. 18% of the SOMMA cohort were classified as having diabetes. The proportion of women to men was greater in the group without diabetes (p=0.02). The proportion of White participants compared to non-White was greater in the group without diabetes (p<0.001). Older adults with diabetes had a higher body weight, waist circumference, BMI, and HbA1c (all, p<0.001). Individuals with diabetes tended to have a higher prevalence of heart disease and hypertension, whereas those without diabetes more often had history of cancer diagnosis (p<0.001). Approximately 75% of participants with diabetes reported use of a hypoglycemic agent where 83% used metformin 19% used insulin, and 2% used thiazolidinediones (TZD). Total daily steps, and time in MVPA was significantly lower (p<0.001 for both) and total sedentary time was significantly higher in those with diabetes (p<0.001). Quadriceps muscle fat infiltration (QFI) was higher in those with diabetes (p=0.002). In addition, both abdominal subcutaneous adipose tissue (ASAT) and visceral adipose tissue (VAT) volume was significantly higher in older adults with diabetes (p=0.005 for ASAT and p<0.001 for VAT) (**Table 1**).

**Table 1:** Participant Characteristics.

| Basic Characteristics | Without Diabetes | With Diabetes | p-value |
| --- | --- | --- | --- |
| N | 717 | 159 |  |
| Age (year) | 76.47 ± 5.05 | 75.79 ± 4.84 | 0.12 |
| Female | 61(438) | 51(81) | **0.02** |
| Race (white) | 88(631) | 70(111) | **<0.001** |
| Hispanic ethnicity | 1(7) | 1(2) | 0.76 |
| Weight (kg) | 74.74 ± 14.75 | 82.41 ± 15.58 | **<0.001** |
| Height (m) | 1.66 ± 0.10 | 1.67 ± 0.10 | 0.16 |
| Waist circumference (cm) | 92.69 ± 12.98 | 100.90 ± 12.57 | **<0.001** |
| BMI (kg/m<sup>2</sup>) | 27.17 ± 4.44 | 29.55 ± 4.63 | **<0.001** |
| HbA1c (%) | 5.51 ± 0.31 | 6.73 ± 0.89 | **<0.001** |
| Systolic blood pressure (mmHg) | 129.71 ± 15.80 | 133.72 ± 16.08 | **0.004** |
| **Co-morbidities** |  |  |  |
| Lung disease | 13(93) | 14(22) | 0.77 |
| Arthritis | 57(409) | 52 (83) | 0.25 |
| Stroke | 2(16) | 3(5) | 0.51 |
| Heart disease | 6(42) | 11(18) | **0.02** |
| Cancer | 43(308) | 24(38) | **<0.001** |
| Hypertension | 26(187) | 37(59) | **0.006** |
| Kidney disease/renal failure | 3(24) | 6(9) | 0.19 |
| Peripheral vascular disease | 1(4) | 1(2) | 0.37 |
| Hypothyroidism | 19(133) | 17(27) | 0.64 |
| Glaucoma | 9(65) | 11(18) | 0.39 |
| Macular degeneration | 9(61) | 5(8) | 0.12 |
| Cataracts | 66(472) | 62(99) | 0.37 |
| **Hypoglycemic Medications** |  |  |  |
| All | -- | 75(118) |  |
| • Insulin | -- | 19(23) |  |
| • Metformin | -- | 84(99) |  |
| • Thiazolidinediones | -- | 3(3) |  |
| **Physical Activity** |  |  |  |
| Total daily steps | 7066.68 ± 3233.95 | 5848.70 ± 2791.12 | **<0.001** |
| Total sedentary time (min) | 610.10 ± 112.28 | 659.40 ± 108.65 | **<0.001** |
| Time in light physical activity (min) | 98.20 ± 26.54 | 92.84 ± 29.16 | **0.027** |
| Time in moderate to vigorous physical activity (min) | 192.95 ± 86.40 | 155.66 ± 76.74 | **<0.001** |
| **Body Adiposity** |  |  |  |
| Abdominal adipose tissue volume (L) | 7.50 ± 3.14 | 8.30 ± 3.31 | **0.005** |
| Visceral adipose tissue volume (L) | 3.96 ± 2.21 | 5.36 ± 2.31 | **<0.001** |
| Quadri-cep fat infiltration (%) | 0.07 ± 0.02 | 0.07 ± 0.02 | **0.002** |

Data presented as means ± standard deviation or % (N). Hypoglycemic medications is calculated only for those taking medications. Significance accepted at **p<0.05** and indicated with bold font.

### Cardiorespiratory Fitness and Muscle Energetics in Diabetes

In the adjusted model (**Table 2**, model 1 + PA), we report that VO_2_peak was significantly lower in older adults with diabetes compared to those without (p= 0.004). State 3 (coupled) respiration in the presence of pyruvate + malate (OXPHOS_CHO_, p= 0.12,) and with the addition of both glutamate and succinate (maxOXPHOS_CHO_, p= 0.10) was similar between groups. Maximal electron transfer system (ETS) capacity measured by maximal uncoupled respiration with CHO substrates (maxETS_CHO_, p=0.03) was significantly lower in those with diabetes (**Table 2**). State 3 (coupled) respiration in the presence of palmitoyl carnitine + malate was similar between those with and without diabetes (OXPHOS_FAO_, p= 0.10). Maximal oxidative phosphorylation supported by fatty acid substrates (maxOXPHOS_FAO_, p=0.006) remained significantly lower in older adults with diabetes in model 1 + PA. Sensitivity analyses completed in those with data for both Protocol 1 and Protocol 2 had similar results (data not shown) to ensure that differences between protocols were not due to differences in sample size.

**Table 2.** Cardiorespiratory fitness and mitochondrial energetics in older adults with and without diabetes.

| Model | Diabetes Status | VO<sub>2</sub>peak | OXPHOS<sub>CHO</sub> | max OXPHOS<sub>CHO</sub> | max ETS<sub>CHO</sub> | OXPHOS<sub>FAO</sub> | max OXPHOS<sub>FAO</sub> |
| --- | --- | --- | --- | --- | --- | --- | --- |
| Unadjusted | WOD | 1532.10 ± 16.67 | **31.08 ± 0.45** | **60.63 ± 0.74** | **81.43 ± 0.97** | 12.94 ± 0.22 | **58.22 ± 0.84** |
|  | WD | 1513.75 ± 36.19 | **28.10 ± 0.97<sup>#</sup>** | **55.11 ± 1.61<sup>#</sup>** | **73.80 ± 2.11<sup>#</sup>** | 11.93 ± 0.47 | **53.31 ± 1.78<sup>#</sup>** |
| Model 1 | WOD | **1486.68 ± 19.07** | **30.14 ± 0.91** | **59.02 ± 1.45** | **77.53 ± 1.90** | **12.58 ± 0.47** | **59.25 ± 1.64** |
|  | WD | **1386.69 ± 25.66<sup>\*</sup>** | **27.88 ± 1.11<sup>#</sup>** | **54.98 ± 1.80<sup>#</sup>** | **71.25 ± 2.37<sup>#</sup>** | **11.49 ± 0.55<sup>#</sup>** | **54.12 ± 1.93<sup>#</sup>** |
| Model 1 + PA | WOD | **1487.30 ± 19.53** | 30.15 ± 0.91 | 58.65 ± 1.47 | **77.02 ± 1.98** | 12.60 ± 0.50 | **59.21 ± 1.61** |
|  | WD | **1412.11 ± 26.03<sup>#</sup>** | 28.51 ± 1.11 | 55.89 ± 1.78 | **77.83 ± 2.44<sup>#</sup>** | 11.70 ± 0.59 | **54.29 ± 1.92<sup>#</sup>** |
| Model 1 + ASAT | WOD | **1492.37 ± 19.52** | **30.34 ± 0.94** | **59.50 ± 1.49** | **77.64 ± 1.95** | **12.67 ± 0.48** | **59.83 ± 1.67** |
|  | WD | **1376.07 ± 26.56<sup>\*</sup>** | **27.83 ± 1.15<sup>#</sup>** | **55.07 ± 1.82<sup>#</sup>** | **70.92 ± 2.45<sup>#</sup>** | **11.37 ± 0.57<sup>#</sup>** | **53.85 ± 1.98<sup>#</sup>** |
| Model 1 + VAT | WOD | **1488.92 ± 19.56** | 30.16 ± 0.94 | **59.16 ± 1.49** | **77.33 ± 1.96** | **12.62 ± 0.49** | **59.57 ± 1.68** |
|  | WD | **1386.86 ± 26.60<sup>\*</sup>** | 28.04 ± 1.15 | **55.46 ± 1.81<sup>#</sup>** | **71.29 ± 2.45<sup>#</sup>** | **11.50 ± 0.57<sup>#</sup>** | **54.66 ± 1.98<sup>#</sup>** |
| Model 1 + QFI | WOD | **1486.37 ± 19.00** | **30.28 ± 0.93** | **59.38 ± 1.48** | **77.78 ± 1.94** | **12.68 ± 0.48** | **59.74 ± 1.68** |
|  | WD | **1386.10 ± 25.75<sup>\*</sup>** | **27.97 ± 1.14<sup>#</sup>** | **55.33 ± 1.81<sup>#</sup>** | **71.06 ± 2.43<sup>#</sup>** | **11.37 ± 0.57<sup>#</sup>** | **54.22 ± 1.98<sup>#</sup>** |

Mean ± standard error with significance accepted at \***p<0.001**, #**p<0.05** and represented with bold font. Model 1 is adjusted for age, race, gender, site/technician, BMI, and comorbidities. PA = physical activity, ASAT = abdominal subcutaneous adipose tissue, VAT = visceral adipose tissue, QFI = muscle quadricep fat infiltration, WOD = without diabetes, WD = with diabetes, OXPHOS<sub>CHO</sub> = carbohydrate supported respiration, maxOXPHOS<sub>CHO</sub> = carbohydrate supported maximal complex I+II stimulated respiration, maxETS<sub>CHO</sub> = carbohydrate supported maximal electron transport system, OXPHOS<sub>FAO</sub> = fatty acid oxidation (FAO) supported respiration, maxOXPHOS<sub>FAO</sub> = fatty acid oxidation supported maximal complex I+II stimulated respiration.

Adjustment for levels of QFI, ASAT and VAT did not impact group differences in mitochondrial energetics and VO_2_peak measurements with an exception for OXPHOS_CHO_ (**Table 2**). Adjustment for VAT tended to explain some variance in OXPHOS_CHO_ between those with and without diabetes (p=0.06, **Table 2**). Within the diabetes group only, ATPmax was lower in older adults taking any hypoglycemic medication (p=0.05) or metformin alone (p=0.01, **Supplemental table 1**). These findings make it difficult to distinguish the impact of medications versus diabetes on ATPmax and therefore, further analysis of mitochondrial energetics focused on respiration measurements. VO_2_peak and mitochondrial energetics assessed by respiration were similar with the use of all hypoglycemic agents and metformin (**Supplemental Table 1**).

### VO_2_peak and Mitochondrial Energetics Mediate the Relationship between Diabetes and Walking Speed

Lastly, 400-m and 4-m walking speeds were significantly lower in those with diabetes compared to those without diabetes following adjustments for gender, age, and race (base model, **Table 3**). To assess whether VO_2_peak and mitochondrial energetics mediated the relationship between diabetes and walk speed, we compared the β coefficient for mean 400-m and 4-m walking speed from the base linear regression model with the β coefficient of models that included VO_2_peak and/or mitochondrial energetics variables. Additional individual adjustments for maxOXPHOS_CHO_ (400-m, p=0.02, model 2), maxOXPHOS_FAO_ (400-m, p=0.02; model 3) and VO_2_peak (400-m p=0.01; model 4) resulted in no significant change in group differences in 400-m walking speed between those with and those without diabetes (**Table 3**). Alternatively, for 4-m walking speed, further adjustments for maxOXPHOS_CHO_ (4-m, p=0.12; model 2) and maxOXPHOS_FAO_ (4-m, p=0.51, model 3) significantly mediated differences between groups, whereas VO_2_peak (4-m, p=0.04, model 4) did not. Examining the impact of combinations of VO_2_peak and mitochondrial energetics revealed that VO_2_peak with maxOXPHOS_CHO_ (400-m p=0.07, 4-m p=0.27; model 5) or maxOXPHOS_FAO_ (400-m, p=0.20, 4-m, p=0.99) mediated differences between those with and without diabetes. Full adjustments including potential confounders/mediators BMI and chronic conditions explained the remaining variance in walking speed between groups. (**Table 3**). Comparing the β coefficient for mean 400-m and 4-m walking speed from the base linear regression model with the β coefficient of models that included both VO_2_peak and mitochondrial energetics variables revealed that VO_2_peak and mitochondrial energetics explained an additional ∼46-100% of the variance in 400-m and 4-m walking speed between groups (**Figure 1**).

**Table 3:** Multivariable linear regression analysis for the association of diabetes with 400m and 4m walk speed.

| Multivariable model | Without Diabetes | With Diabetes |  |
| --- | --- | --- | --- |
| | Means ± SE | $\beta \pm$ SE | P |
| **400m walk speed (m/sec)** |  |  |  |
| Base Model: adjusted for gender, age and race | 1.06 ± 0.01 | -0.05 ± 0.01 | **0.00** |
| Model 2: base model plus maxOXPHOS<sub>CHO</sub> | 1.06 ± 0.01 | -0.04 ± 0.02 | **0.02** |
| Model 3: base model plus maxOXPHOS<sub>FAO</sub> | 1.06 ± 0.01 | -0.04 ± 0.02 | **0.02** |
| Model 4: base model plus VO<sub>2</sub>peak | 1.07 ± 0.01 | -0.04 ± 0.01 | **0.01** |
| Model 5: base model plus VO<sub>2</sub>peak and maxOXPHOS<sub>CHO</sub> | 1.07 ± 0.01 | -0.03 ± 0.02 | 0.07 |
| Model 6: base model plus VO<sub>2</sub>peak and maxOXPHOS<sub>FAO</sub> | 1.07 ± 0.01 | -0.02 ± 0.02 | 0.20 |
| Fully adjusted: model 5 plus BMI and comorbidities | 1.06 ± 0.01 | 0.003 ± 0.01 | 0.87 |
| Fully adjusted: model 6 plus BMI and comorbidities | 1.06 ± 0.01 | -0.005 ± 0.02 | 0.78 |
| **4m walk speed (m/sec)** |  |  |  |
| Base Model: adjusted for gender, age, and race | 1.05 ± 0.01 | -0.04 ± 0.02 | **0.01** |
| Model 2: base model plus maxOXPHOS<sub>CHO</sub> | 1.04 ± 0.01 | -0.03 ± 0.02 | 0.12 |
| Model 3: base model plus maxOXPHOS<sub>FAO</sub> | 1.05 ± 0.01 | -0.01 ± 0.02 | 0.51 |
| Model 4: base model plus VO<sub>2</sub>peak | 1.05 ± 0.01 | -0.04 ± 0.02 | **0.04** |
| Model 5: base model plus VO<sub>2</sub>peak and maxOXPHOS<sub>CHO</sub> | 1.05 ± 0.01 | -0.02 ± 0.02 | 0.27 |
| Model 6: base model plus VO<sub>2</sub>peak and maxOXPHOS<sub>FAO</sub> | 1.05 ± 0.01 | 0.00 ± 0.02 | 0.99 |
| Fully adjusted: model 5 plus BMI and comorbidities | 1.05 ± 0.01 | 0.003 ± 0.02 | 0.84 |
| Fully adjusted: model 6 plus BMI and comorbidities | 1.05 ± 0.01 | 0.01 ± 0.02 | 0.50 |

Without diabetes, mean ± standard error. With diabetes, $\beta$ estimate ± standard error with significance accepted at \***p<0.05**.
maxOXPHOS<sub>CHO</sub> = carbohydrate supported maximal complex I+II stimulated respiration, maxOXPHOS<sub>FAO</sub> = fatty acid oxidation supported maximal complex I+II stimulated respiration. Models including maxOXPHOS<sub>CHO</sub> and maxOXPHOS<sub>FAO</sub> are adjusted for technician, and models with VO<sub>2</sub>peak are adjusted for site.

**Figure 1.**
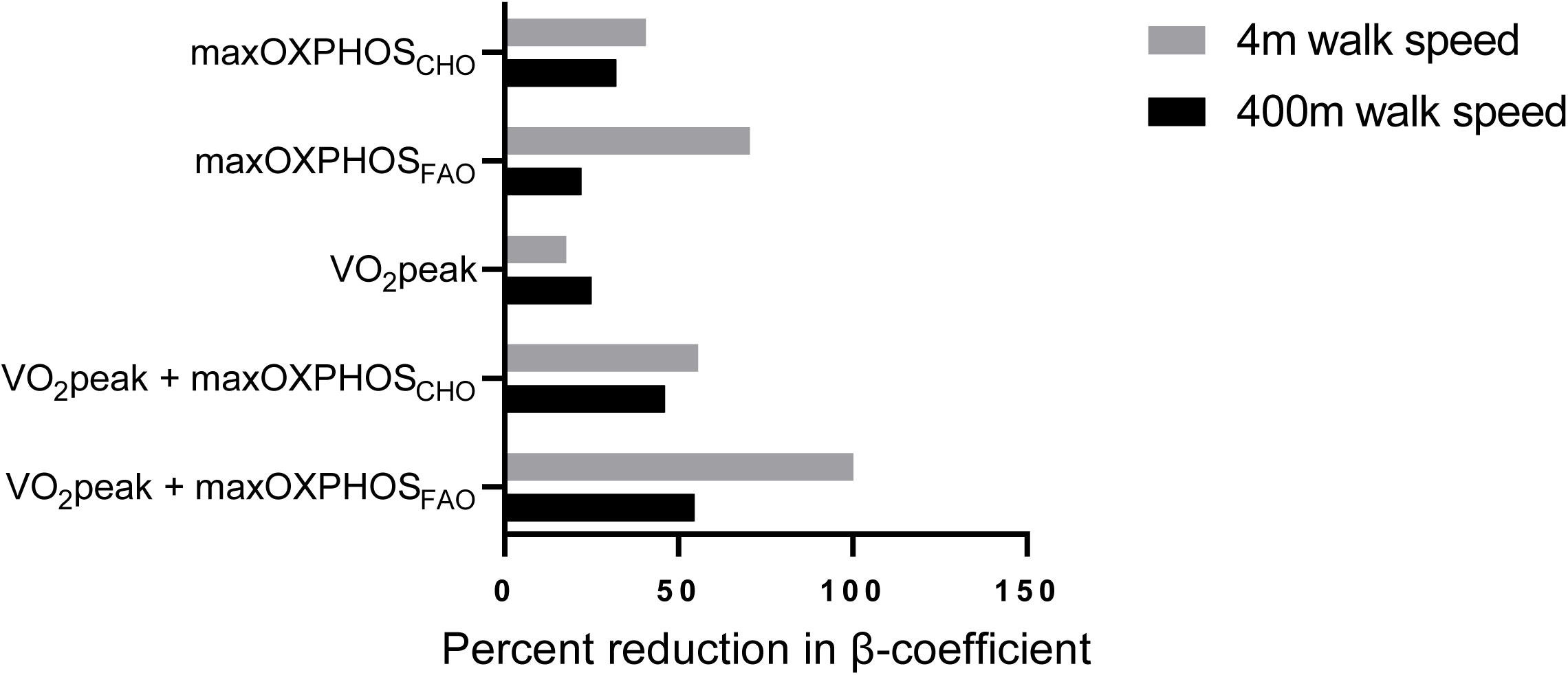
Percent reduction in the association between diabetes and 400-m and 4-m walking speed after adjusting for both individual and combined carbohydrate and fatty acid supported respiration, and cardiorespiratory fitness. Bars depict the reduction in β coefficient from linear regression models (models 2-6 compared to the base model: [1-(β adjusted/β base adjusted)]. Base model: age, gender, and race, model 2: base model + maxOXPHOS_CHO_, model 3: base model + maxOXPHOS_FAO_, model 4: base model + VO_2_peak, model 5: base model + VO2peak + maxOXPHOS_CHO_, model 6: base model + VO_2_peak + maxOXPHOS_FAO_.

## DISCUSSION

In this study of 879 well-phenotyped older adults we found that the 159 participants with diabetes had slower 4- and 400-m walk speed. We report for the first time that the association of diabetes status with slow walking speed was mediated by VO_2_peak and skeletal muscle mitochondrial energetics. We also examined differences in cardiovascular fitness and mitochondrial energetics between those with and without diabetes while controlling for physical activity and adiposity. Interestingly, accounting for these potential confounders did not explain differences in VO_2_peak and mitochondrial respiration between those with and without diabetes, suggesting other factors may impact cardiorespiratory fitness and energetics in those with diabetes. We also found that older adults taking hypoglycemic medications (insulin, TZDs, and/or metformin) or metformin alone had significantly lower ATPmax (but not respirometry measures of mitochondrial energetics) compared to older adults with diabetes but not taking medications, providing evidence that aligns with reports in the literature that metformin can impair mitochondrial energetics.

The well-phenotyped SOMMA cohort provided an opportunity to assess the impact of diabetes status on VO_2_peak and mitochondrial energetics in older adults while controlling for objectively measured indices of physical activity, adiposity, and chronic conditions in addition to demographic variables. Many of the previous studies reporting lower muscle oxidative capacity measured in permeabilized fiber bundles from patients with diabetes have not rigorously assessed and controlled for the participants’ physical activity [12–14, 17]. This is important as physical activity interventions (such as walking) can improve mitochondrial energetics in type 2 diabetes [27] and during weight loss in obesity [28], and in older adults [29]. Here, we report that lower levels of objectively assessed physical activity partially explains lower respiration in older adults with diabetes. Interestingly, comparing our respirometry protocols with distinct substrate combinations, we found that CI+II FAO supported maxOXPHOS respiration remains lower in the diabetes group, while CI+II CHO only supported maxOXPHOS did not differ following adjustments for physical activity. This suggests that CHO supported coupled respiration may be more sensitive to levels of physical activity compared to FAO supported respiration. This is also in line with evidence indicating reduced rates of lipid oxidation in skeletal muscle from individuals with diabetes, potentially due to mitochondrial overload and incomplete fatty acid oxidation [30].

The influence of adiposity on mitochondrial energetics has been considered in prior studies of diabetes, typically by matching control subjects for BMI [14]. Some studies [15, 16], but not all [12, 13, 17], indicate no differences between patients with diabetes and BMI-matched controls, suggesting that obesity *per se* may underlie the lower muscle oxidative capacity. Here, we examined the influence that BMI and specific adipose depots (ASAT, VAT, and QFI) had on muscle mitochondrial energetics in diabetes. However, regardless of whether adiposity was adjusted for BMI alone or combined with individually objectively measured adipose depots, mitochondrial respiration generally remained significantly lower in those with diabetes. Taken together, these findings indicate that adiposity and physical activity largely (there were some exceptions) do not entirely explain lower mitochondrial energetics in older adults with diabetes.

Metformin is one of the most commonly prescribed medications for diabetes, and although its mechanism of action remains highly debated, there is evidence of an inhibitory effect on complex I of the electron transport chain [31]. Both pre-clinical and clinical studies report maladaptation of mitochondrial function following metformin treatment, increased muscle atrophy and reduced exercise capacity, specifically in older adults [31, 32]. Conversely, others have reported that metformin can improve mitochondrial function via effects on mitophagy, autophagy or AMPK activation [33]. Our understanding of how metformin impacts skeletal muscle mitochondrial function of older adults with diabetes is limited. Here, we reveal lower ATPmax in older adults with diabetes taking hypoglycemic medication or metformin alone, compared to older adults with diabetes not taking medications. Interestingly, we did not observe an association of metformin with diabetes status in the respiration assays, suggesting different sensitivities to metformin based on whether mitochondrial energetics are assessed in vivo (ATPmax) compared to permeabilized fiber bundle preparations ex vivo. However, an important caveat is that participants were asked to withhold medication prior to the muscle biopsy and respirometry assays but not on the day of ^31P^MRS assessments. In addition, others have reported no effect on mitochondrial respiration after a two-week metformin treatment compared to control [34], or in long term diabetes patients [35]. Given that metformin is water soluble, it is also plausible that the inhibitory effect of metformin is lost when the muscle fiber bundles are washed in preparation for the respirometry assay. Further work is needed to decipher the impact metformin has on skeletal muscle mitochondrial energetics in older adults with diabetes.

Zhao and colleagues reported that adults with diabetes were 31-34% less likely to participate in physical activity, which in turn may contribute to lower VO_2_peak [36]. In line with these reports, SOMMA participants with diabetes were significantly less active and significantly more sedentary. Our results show that additional adjustments for physical activity did not resolve differences in VO_2_peak between groups, suggesting that lower VO_2_peak in older adults with diabetes is independent of physical activity in this cohort. In addition, we report that independent of adiposity measured by BMI or combined BMI with ASAT, VAT and QFI, VO_2_peak remained significantly lower in those with diabetes compared to those without. Taken together, our work suggests that physical activity and adiposity may not be implicated in lower cardiorespiratory fitness in older adults with diabetes and that other factors including perhaps genetics and heritability are more important determinants to low cardiorespiratory fitness in diabetes.

Previous work from our laboratory and others revealed a significant relationship between walking speed with both VO_2_peak and mitochondrial energetics in older adults [9, 37, 38]. Here, we explored the contribution of both mitochondrial energetics and cardiorespiratory fitness to mobility in older adults with diabetes. We report that mitochondria respiration, both independently (∼40-70%) and combined with VO_2_peak (∼55-100%), mediated the variance in 4-m walking speed in those with diabetes. Differences between groups are further explained by additional adjustments for potential confounders/mediators BMI, and comorbidities. Reports indicate that muscle mass is lost at an accelerated rate in older adults with diabetes [7], contributing to reductions in muscle strength and quality [6], and ultimately reducing physical function and mobility [5, 39]. We extended these findings by highlighting how mitochondrial energetics may independently and combined with cardiorespiratory fitness also contribute to lower walk speed in older adults with diabetes. This finding is of particular interest because slower walking speed is an important indicator of health and has been shown to associate with survival in older adults [38]. This data raises the possibility that cardiorespiratory physiology and mitochondrial energetics could be targeted to prevent mobility loss in older adults with diabetes.

There are some limitations to the study that should be considered. First, the duration of diabetes status of SOMMA participants and the length of time that participants have been taking oral hypoglycemic agents were not recorded. We acknowledge that our study participants were predominantly (85.6%) non-Hispanic White which limits our ability to generalize to other race/ethnic groups where mobility disability is more prevalent [40]. In addition, all participants recruited were able to complete 4-m walk test at a gait speed of greater than or equal to 0.6m/s and which may be faster than those with more advanced diabetes who are unable to complete mobility related tasks [2]. Nevertheless, we observe differences in physiological and functional aspects of skeletal muscle signifying the impact that diabetes has in the SOMMA cohort. The strengths of our study are the large sample size of participants and rigorous assessment of objectively measured physical activity, fitness and body composition, and collection of muscle biopsies for fresh tissue assays.

In summary, our findings highlight that mitochondrial energetics independently and combined with cardiorespiratory fitness contribute to the difference in 400-m and 4-m walk speed in older adults with diabetes. Additionally, future work should aim to decipher the impact of diabetes medication on mitochondrial function as this remains to be a gap in the literature.

## Data Availability

All Data produced in the present study are available upon reasonable request to the authors

## Personal thanks

We gratefully acknowledge and thank all participants who participated in this study at the University of Pittsburgh and Wake Forest University. We acknowledge the dedicated staff and investigators at both clinical sites as well as at the San Francisco Coordinating Center. Collection and analyses of ^31^Phosphorous Magnetic Spectroscopy data would not have been possible without the support from Eric Shankland.

## Funding and Assistance

The National Institute on Aging (NIA) funded the Study of Muscle, Mobility, and Aging (SOMMA; R01AG059416). In part, infrastructure support for SOMMA was funded by the NIA Claude D. Pepper Older American Independence Centers at the University of Pittsburgh (Pitt) and Wake Forest University School of Medicine (Wake), P30AG024827 and P30AG021332 respectively. More SOMMA infrastructure support from the Clinical and Translational Science Institutes is funded by the National Center for Advancing Translational Science at both Wake (UL1TR001420). PMCo is supported by R01AG060153 and R01AG060542. GD was supported by the American Diabetes Association (1-19-PDF-006) during data collection and analysis.

## Author Contributions and Guarantor Statement

SVR and LYL completed statistical analysis with final review from the statistical team at the coordinating center. SVR and PMCo conceived the idea and co-wrote the manuscript. LYL, GD, SM, PMCo and PK completed experiments or completed quality control, validation, and interpretation of data. LYL, GD, PMCa, PK, TM, SM, MJJ, AJM, EEK, DJM, FGST, SRC, ABN, RTH, SBK, BHG edited the manuscript. SRC, PMCa, ABN, SBK, RTH, and BHG enabled the study with either funding acquisition, project administration, and/or conceptualization to the study.

## Prior Presentations

Data from this manuscript has been presented at the following meetings: American Diabetes Association 83^rd^ Scientific Sessions May 30^th^ - June 6^th^ 2022, and the American College of Sports Medicine Annual Meeting and World Congress, May 30^th^ – June 2^nd^, 2023.

---

**Supplemental Figure 1.**
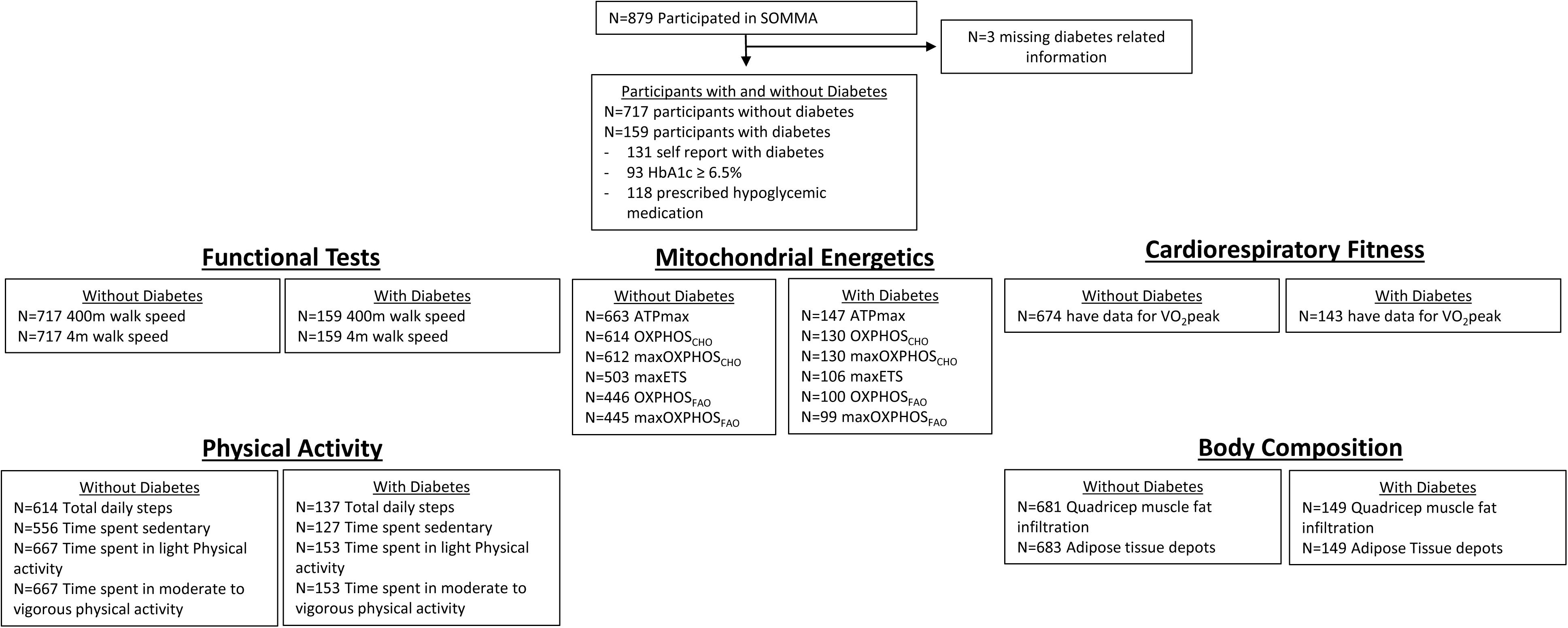
SOMMA cohort sample size for analysis of muscle function, mitochondrial energetics, CRF, PA and body composition. Participants missing data for ex-vivo mitochondrial energetics is due exclusions samples passing quality control. Missing ATPmax measures were due to safety reasons or exclusion due to technical conditions during testing.

**Supplemental Table 1.** Association of hypoglycemic drugs and metformin with cardiorespiratory fitness and mitochondrial energetics.

|  | All hypoglycemic agents |  | Metformin |  |
| --- | --- | --- | --- | --- |
| | $\beta$ estimate | p-value | $\beta$ estimate | p-value |
| VO<sub>2</sub>peak | -82.27 ± 62.24 | 0.19 | -13.08 ± 55.47 | 0.81 |
| ATPmax | **-0.05 ± 0.03** | **0.05** | **-0.06 ± 0.02** | **0.01\*** |
| OXPHOS<sub>CHO</sub> | -1.03 ± 2.01 | 0.61 | -0.71 ± 1.81 | 0.70 |
| maxOXPHOS<sub>CHO</sub> | -3.52 ± 3.20 | 0.27 | -3.11 ± 2.89 | 0.28 |
| maxETS<sub>CHO</sub> | -2.95 ± 4.45 | 0.51 | -4.07 ± 4.04 | 0.32 |
| OXPHOS<sub>FAO</sub> | -1.35 ± 1.06 | 0.21 | -0.68 ± 1.00 | 0.50 |
| maxOXPHOS<sub>FAO</sub> | -2.57 ± 3.69 | 0.49 | -5.30 ± 3.43 | 0.13 |

$\beta$ estimate ± standard error with significance accepted at **p<0.05**. Data adjusted for age, race, and site/technician. ATPmax = maximal adenosine triphosphate production, OXPHOS<sub>CHO</sub> = carbohydrate supported respiration, maxOXPHOS<sub>CHO</sub> = carbohydrate supported maximal complex I+II stimulated respiration, maxETS<sub>CHO</sub> = carbohydrate supported maximal electron transport system, OXPHOS<sub>FAO</sub> = fatty acid oxidation (FAO) supported respiration, maxOXPHOS<sub>FAO</sub> = fatty acid oxidation supported maximal complex I+II stimulated respiration.

